# Non-technical skills in Obstetric Aeromedical Transfers (NOAT): development and evaluation of a behavioural marker system

**DOI:** 10.1101/2022.08.29.22279364

**Authors:** Anuradha Perera, Julia A. Myers, Robin F Griffiths

## Abstract

**Background:** Non-technical skills contribute to patient safety and multidisciplinary team performance in acute and complex health care settings. Behavioural frameworks are increasingly being used in health care to teach and evaluate non-technical skills. A framework specific to the maternity aeromedical transfer setting would be highly beneficial, because of the potential impact of non-technical skills on patient outcomes in this highly complex and dynamic clinical setting.

**Methods:** Taking a transformative approach, an existing non-technical skills framework was adapted to the maternity aeromedical transfer setting. Initially, non-technical skills and behavioural markers specific to maternity aeromedical transfer were identified by triangulating data from focus group interviews, field observations, and the literature. Content analysis was used to code and sort data to develop the Non-technical skills in Obstetric Aeromedical Transfers (NOAT) framework. Finally, we evaluated the NOAT prototype for its basic psychometric properties such as feasibility, face validity and content validity by surveying clinicians experienced in maternity aeromedical transfers, direct field observations, and clinical simulation.

**Results:** The NOAT framework consists of six main skills categories:

- Communication with the patient and partner,
- Task/case management,
- Teamwork,
- Situational awareness,
- Communication with team members,
- Environment of the cabin.

A rating scale can be used to assign numerical values to non-technical skill performance and emphasise the relevance to patient safety. Initial evaluation indicates the basic psychometric properties of NOAT including feasibility of use, content validity and face validity are positive.

**Conclusion:** The NOAT framework identifies specific non-technical skills and their behaviours applied to a maternity aeromedical transfer setting. It has the potential to provide a framework around which teaching, training, and debriefing can be structured. Integration of non-technical skills competency training has the potential to significantly enhance the safety of women being transported in high acuity situations.

**Key messages:** *What is already known on this topic?:*

➢ A validated framework can provide a common language for comprehensively and consistently measuring non-technical skills specific to the clinical setting in which it is applied.

*What this study adds:*

➢ The Non-technical skills in Obstetric Aeromedical Transfers (NOAT) framework provides a valid and reliable system for the application and training of non-technical skills in the highly complex and dynamic maternity aeromedical transfer setting.

*How this study might affect research, practice or policy:*

➢ Applying a non-technical skills framework based on potential patient safety hazards provides an ideal opportunity to promote the safety of pregnant women being transported in high acuity clinical situations.
➢ The NOAT framework can be used as a reference point for maternity aeromedical transfer teams’ in-service programmes, and for service managers and policy developers to make evidenced-based decisions for organisational enhancements in this clinical setting.

## Introduction

### Adverse Events in Health Care

Adverse events are more prevalent in acute and complex areas of health care, posing a major threat to patient safety and quality of care (1–6). In addition to high levels of technical expertise, the avoidance of adverse effects in these clinical settings frequently requires effective communication between multidisciplinary team members, and coordination of multiple parallel activities with high levels of situational awareness (7–10). The importance of non-technical skills has become increasingly recognised in health care (11–13). The term non-technical skills is defined as ‘the cognitive, social, and personal resource skills that complement technical skills, and contribute to safe and efficient task performance’ (14).

Comprehensive application of non-technical skills across health care settings provides opportunities to provide additional protection for patients (15–18). The risk of adverse events due to poor non-technical skill performance is not unique to one particular health speciality; rather, it is a global concern for the overall safety and quality of health care delivery (5, 19–24). Non-technical skills like teamwork, communication, situational awareness, task management, decision making and leadership are recognised by the World Health Organization as key to preventing, recognising, and mitigating errors, making them critical to patient safety (25, 26)

### Relevance of Non-technical Skills in the Maternity Aeromedical Transfer Setting

An increasing body of knowledge in maternity care supports the view that non-technical skills are paramount for maintaining safe clinical performance (27–33). For example, a recent report from the Royal College of Obstetricians and Gynaecologists identified situational awareness to be a factor in 44%, intra/interprofessional communication in 45%, and team leadership in 22% of adverse events and patient harms (34). This evidence demonstrates why clinicians’ awareness of how non-technical skills and behaviours can be purposefully employed is so important in defending against avoidable human errors in maternity settings (31, 35, 36).

Transfer of high-acuity obstetric patients by air ambulance from provincial maternity units to tertiary facilities demonstrates a significant benefit to neonatal and maternal outcomes. This is particularly important for countries with dispersedpopulations where highly specialised maternity care is more centralised, making timely access critical (37–45). Maternity aeromedical transfers are usually staffed by aeromedically trained health professionals and mostly undertaken in fixed-wing aircraft, while a helicopter is used in more urgent situations (44, 46). The transfer team consists of a midwife, and a flight nurse or paramedic on most occasions, while a doctor is on board occasionally, depending on the clinical complexity or acuity of the pregnant woman (45).

In this context, in addition to acute clinical care required for the pregnant woman and foetus, there are aviation-related factors such as noise, vibration, low atmospheric pressure, poor lighting, and space constraints that further challenge patient safety (45, 47, 48). For these reasons, clinicians and researchers must understand exactly which non-technical skills are specifically applicable to ensure the safety of a pregnant woman when transported by air ambulance (49).

To provide the basis for measuring the non-technical skills performance of clinicians or health care teams, non-technical skills frameworks specific to health care settings have been developed over the last two decades (50–53). These non-technical skills frameworks are often referred to as ‘behavioural marker systems’, as they are based on pre-defined behaviours that can be observed by an independent observer (14, 54). These behaviours are specific to a clinical setting (55–57). Non-technical skills frameworks assist the teaching and assessment of clinical performance, and provide a common structure and language for promoting non-technical skills (11, 58, 59). The necessity of developing setting specific strategies for maximising the non-technical skills performance of health professionals and promoting safe and reliable health care delivery is now well recognised (60, 61). However, a context-specific framework to evaluate non-technical skills in a maternity aeromedical transfer setting was not available prior to this study.

### Study Aims

The present study aimed to address this gap by developing a valid and feasible framework for observation-based rating and constructive debriefing of the non-technical skills performance of maternity aeromedical transfer teams.

## Methodology

Non-technical skills categories are largely generic; what varies from one health context to the next are the individual skill elements and behaviours (behavioural markers) clinicians display in demonstrating the non-technical skills (14). Based on an integrative review of existing non-technical skills frameworks, the Global Assessment of Obstetric Team Performance (GAOTP) framework was selected as the most suitable framework to adapt to the aeromedical maternity transfer environment (49). The rationale for this selection was that GAOTP was designed for an obstetric clinical setting, is team-focused, and has been shown to be effective for rating multidisciplinary obstetric teams’ non-technical skills performance in hospital-based delivery rooms and theatre contexts (62, 63). Using GAOTP as the blueprint, we then followed recommended approaches from the literature to develop our initial prototype of the Non-technical skills in Obstetric Aeromedical Transfers (NOAT) framework. In particular, the recommendations mandate that when an existing framework is adapted to a different clinical setting, the work must be supplemented by data collected directly from the clinical setting to which the framework is being applied, ensuring the identification of setting-specific behaviours (64).

The development and preliminary evaluation of the NOAT framework involved two phases. In the first phase, the NOAT framework was developed by triangulating data from health professionals involved in maternity aeromedical transfers, literature-based evidence, and direct field observation. Content analysis was able to identify unique non-technical skill behaviours determinative of maternity aeromedical team functioning as part of a coherent framework (65). In the second phase, the basic psychometric properties of the NOAT framework were evaluated based on clinicians’ experiences in maternity aeromedical transfer, direct field observations, and clinical simulation.

### Interviews with key stakeholders

We purposively sampled and interviewed 12 data-rich subject matter experts (service managers and clinicians) representing various District Health Boards (DHBs) across New Zealand, using semi-structured questions. If any issues remained unanswered at the end of this process, some additional questions or clarifications were used to capture that information (66).

### Direct field observations

Direct field observation was conducted with a regional air ambulance service. Observations provided details of observed behaviours exhibited by maternity aeromedical transfer teams.

### Survey

Qualtrics electronic survey software enabled the development and generation of an anonymous online survey distributed via DHBs and professional organisations to flight midwives, flight nurses, and paramedics who routinely perform maternity aeromedical transfers. We used a five-point scale from ‘1-not at all important’ up to ‘5-essential’ in the survey, for respondents to rate the importance of each skill. An additional comments section was available for clinicians to suggest skills or behaviours they believed were important, but not included (Supplementary document 1).

### Clinical simulation study

A simulation-based training exercise provided further contributory data (67–69). Midwives and flight nurses undertaking the training provided informed consent to have their case scenarios video recorded. The clinical scenarios recorded were based on potential maternity aeromedical emergencies. The research team later independently reviewed the scenario recordings, applying the NOAT framework to rate all non-technical skills they observed during the scenario. The NOAT framework was assessed on its ability to adequately capture any non-technical skills observed during those scenarios (70).

## Analysis

Deductive content analysis based on the core non-technical skills categories of the GAOTP framework were used to identify unique non-technical skill behaviours from interview transcripts, and field based observations (65). These behaviours were organised according to the six skills categories of the NOAT framework. The resulting prototype was then refined and consolidated iteratively by making further observations in the maternity transfer environment, and by re-coding a sample of data. We used the following recommended criteria when developing the NOAT behavioural marker system (71):

- the skills and their behaviours were observable
- there was a hierarchical structure to the framework
- the NOAT framework could be used with minimal training.

We calculated the Content Validity Index (CVI) of the behaviours based on the survey results, by dividing the total number of respondents into the number of participants who rated the various behavioural descriptors at 4 or 5 (‘very important’ or ‘essential’). We reviewed and revised any behavioural descriptors not reaching a level of at least 0.75 (72). Data from the free text questions were also analysed and categorised to determine whether they provided any new skill categories or behavioural markers (72).

## Results

### The NOAT framework

The final NOAT framework consists of the six main overarching non-technical skills categories from the GAOTP framework (Table 1):

- Communication with the pregnant woman and the support person/family
- Task management
- Teamwork
- Situational awareness
- Communication with team members
- The environment of the cabin.

**Table 1:**
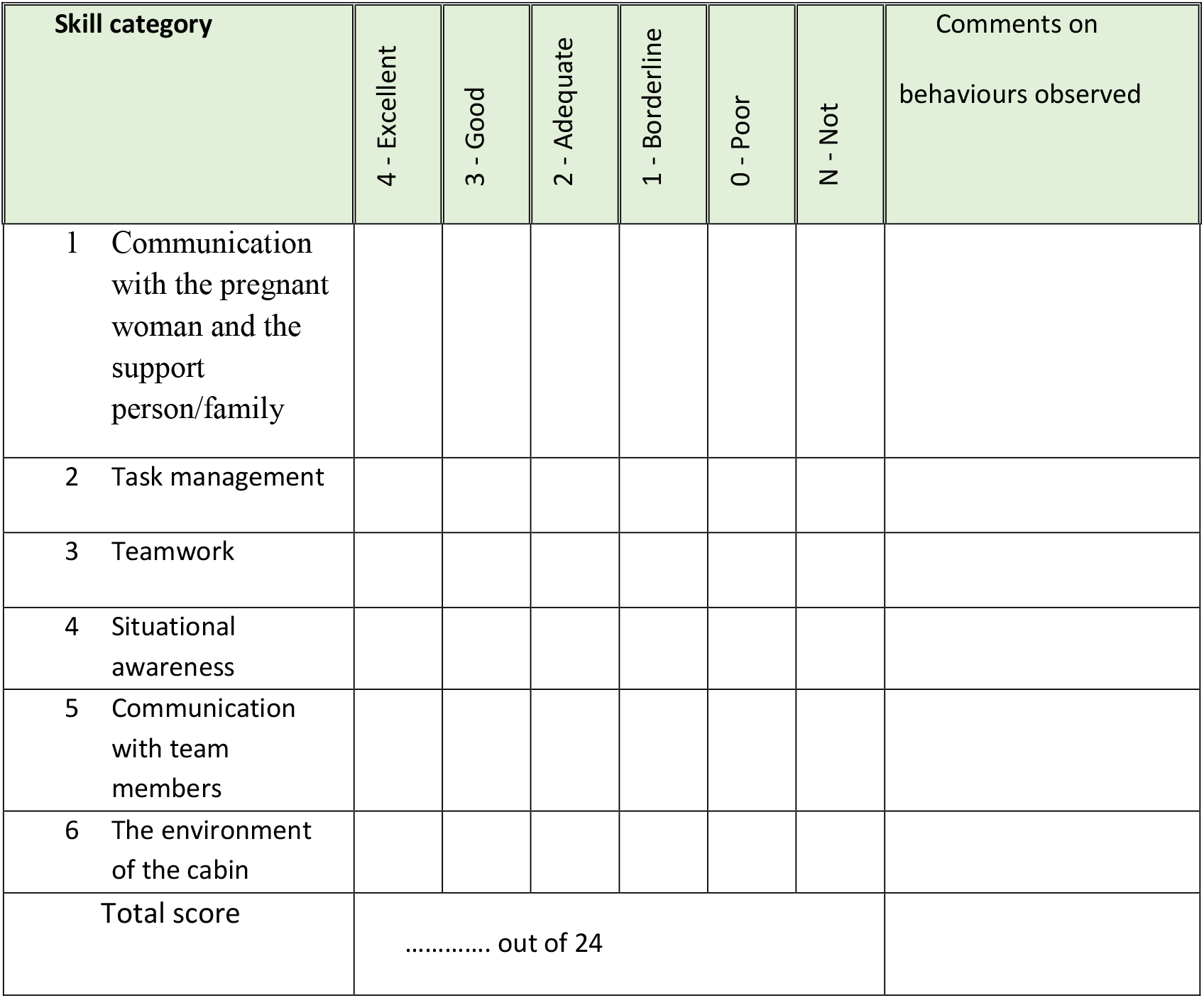
The Non-technical skills in Obstetric Aeromedical Transfers (NOAT) framework.

A 5-point Likert scale and a space to write brief comments is also included. The rating scale enables assessors to assign numbered scores based on the quality or presence of observed behaviours (Table 2). For example, the highest rating of ‘4’ equates to ‘Excellent’, indicating an extremely good performance, considered as best practice and serving as a model example for others. A rating of ‘0’ indicates a ‘Poor’ performance with the absence of the required behaviour, and a performance which endangered or potentially endangered patient safety. We also suggest an additional rating option of ‘Not required’ (N) when the skill is not needed in a particular situation. Each skill category of the NOAT framework elaborated below has a definition and exemplar behavioural markers, which were identified and supported by quotations from field observations and subject matter expert interviews, and triangulated with the evidence from scientific literature (Table 3).

**Table 2:**
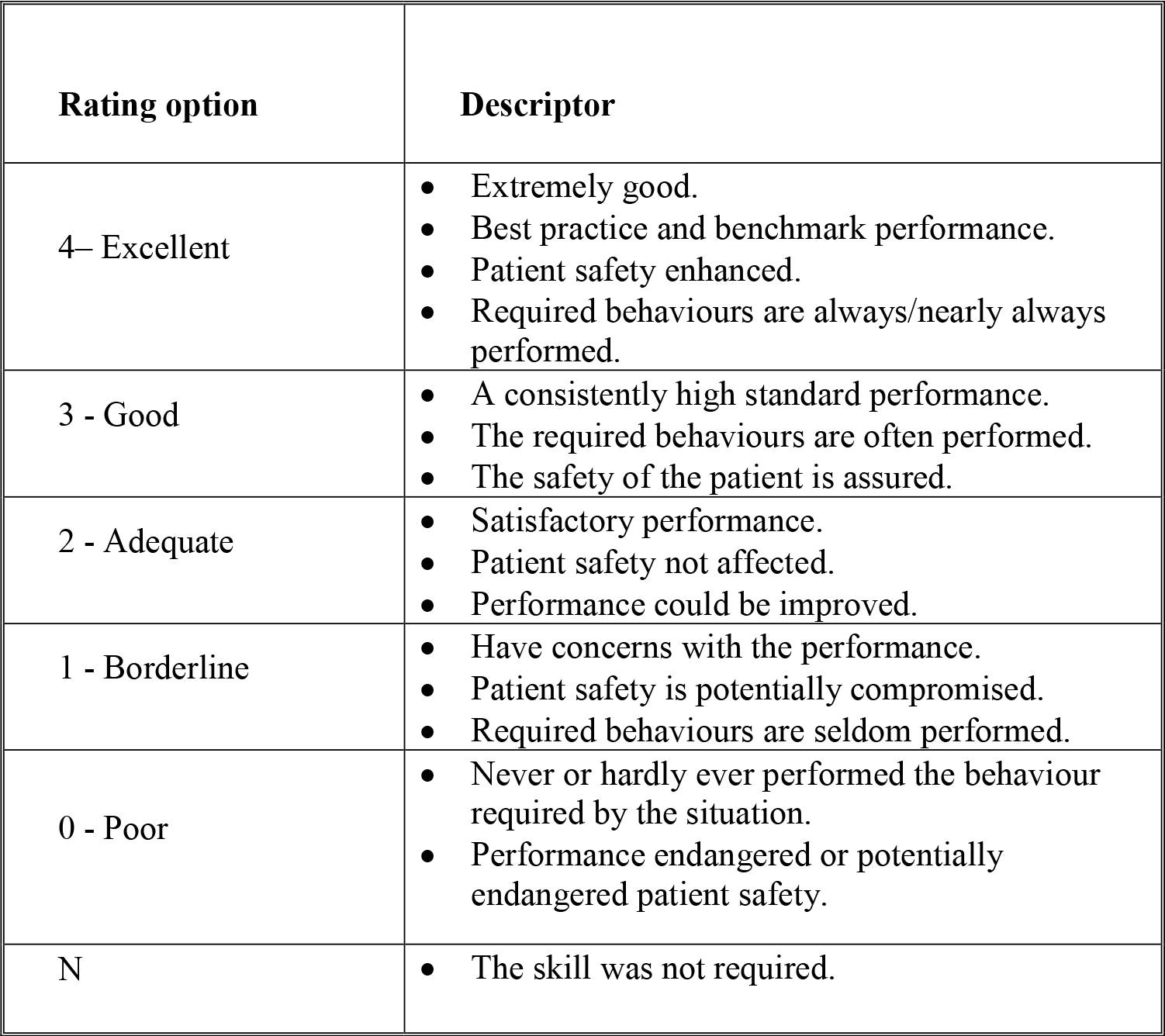
Performance descriptors of the rating scale.

**Table 3:** Good exemplar behaviours of skill categories in the NOAT framework.

|  |
| --- |
| **1 Communication with the pregnant woman and the support person/family** |
| • Making introductions to the woman and partner/family member and identifying team roles when you take over their care. |
| • Checking the understanding and ability of the woman and her partner/family member to be involved in decisions over care. |
| • Active and continuous information sharing and involvement of the woman/partner/family in care decisions. |
| • At least one team member assumes the responsibility for continuously reassuring the woman/partner/family during transfer. |
| • Demonstrating empathy for the situation by encouraging the woman and her partner/family member to verbalise their thoughts and feelings. |
| • Demonstrating the ability to elicit relevant information from the woman and her partner/family member while appreciating their concerns and emotions. |
| • Practicing assertive but not intimidating behaviour with the woman/partner/family. |
| • Performing a pre-flight briefing for the woman and her partner/family member. |
| • Taking active measures to avoid or dispel the partner's/family member's disruptive response to a changing clinical situation. |
| **2 Task management** |
| • Organising and negotiating ahead of time for an appropriate plan for transport and arrival of the woman with relevant parties. |
| • Sourcing relevant information from the referral hospital. |
| • Developing a plan of care between team members and other relevant parties. |
| • Collecting all relevant information and objectively considering a go/no go decision on the flight. |
| • Cross-checking all medications and other equipment before departure. |
| • Considering the woman's care needs when exploring transport options (i.e. fixed-wing or helicopter). |
| • Considering the urgency and priorities of the situation when making or reviewing the plan. |
| • Preparing the woman ahead of time for procedures. |
| • Verifying with team members before proceeding with interventions. |
| • Following established protocols and guidelines. |
| • Maintaining accurate transport records. |
| • Identifying all resources that are available and how they operate. |
| • Delegating tasks to others when required. |
| **3 Teamwork** |
| • Establishing roles with clear and confirmed responsibilities. |
| • Working together to carry out tasks with a shared understanding. |
| • Demonstrating trust and respect to the team members for their abilities. |
| • Avoiding criticising. |
| • Encouraging team members to give their opinions and demonstrating active listening skills. |
| • Team members providing back-up for each other. |
| • Team members openly accepting help. |
| • Considering team composition when planning the mission. |
| • Taking over leadership when required. |
| • Questioning another team member when unsure about a decision they have made. |
| • Team members sharing their own expertise, capabilities, and limitations. |
| • Providing encouragement/reassurance to colleagues. |
| • Empowering team members to report incidents. |
| **4 Situational Awareness** |
| • Team members remaining vigilant and alert to the clinical situation. |
| • Gathering necessary documents before the transport. |
| • Frequently monitoring the woman and the environment. |
| • Understanding the aviation/transport-related risks to the woman/family. |
| • Collecting and sharing information from the team members and woman to understand the situation. |
| • Continuously recognising all possible alterations as the woman's condition evolves. |
| • Recognising early on and responding rapidly to critical/ changing situations. |
| Adapting to changing situations. |
| Recognising each other's limitations relating to knowledge, skills, and ability and taking pre-emptive actions. |
| Planning ahead of time for interventions and understanding possible limitations for interventions due to the restrictive airborne setting. |
| Anticipating problems and taking actions to mitigate them. |
| Continuous planning and preparing for the next phase of the mission. |
| Reviewing the situation after key stages of the transfer. |
| **5 Communication with Team Members** |
| Focused communication with clear questions/instructions directed toward the other members. |
| Regularly updating each other with relevant information. |
| Reaching mutual understanding by not only exchanging information but confirming a shared understanding. |
| Using a closed-loop communication technique to communicate. |
| Communicating and coordinating the woman's interventions with the flight deck. |
| Conducting a formal pre-flight briefing. |
| Verbalising own observations and their meaning to other team member/s. |
| Cross-checking all information. |
| Acknowledging concerns to others. |
| Ensuring proper handover and takeovers. |
| Speaking up when mistakes are made and making suggestions without fear of negative consequences. |
| Conducting formal debriefing sessions. |
| Reporting incidents as a part of the transfer. |
| **6 Environment in the Cabin** |
| Voices and body language remain calm and focused on the clinical situation. |
| Orderly and controlled cabin. |
| Demeanour is controlled. |
| • Maintaining calm, confident, caring, and reassuring behaviour with woman/partner/family member. |
| • Demonstrating an understanding of the clinical and unfamiliar nature of care to the woman/partner/family member. |
| • Helping to ease the woman's anxiety and aid the transition from a personalised labour room to an aviation-based clinical environment. |
| • Maintaining professional behaviour. |
| • Maintaining the dignity of the woman and her partner/family member |

#### Communication with the Patient and the Partner

In maternity aeromedical transfers, clinicians are caring for an essentially well, conscious pregnant woman in a dynamic environment. Subject matter experts who participated in this study reported that the non-technical skill of developing a professional relationship with the pregnant woman and her partner in a time-limited and stressful situation is crucial to both the patient experience and clinical outcomes. This includes appreciating a woman’s potential cultural and social orientation, respect and dignity when communicating, demonstrating empathy, acknowledging their distress, and providing continuous psychological support (73).

Unlike other aeromedical transfers, there is a high degree of partner and family involvement, posing communication and management challenges for the clinical team. Study participants recommended that health professionals should practice assertive behaviours, but should take care not to intimidate (Table 4).

**Table 4:** Sample quotations from subject matter expert interviews and field notes to support the each skill category.

| Communication<br>with the patient<br>and the partner | Task management | Teamwork | Situational awareness | Communication with<br>team members | The environment of the<br>cabin. |
| --- | --- | --- | --- | --- | --- |
| “All of a sudden you have to give them (family) a familiarisation of the aircraft, a safety briefing., in the middle of the night straight on a stretcher, headphones on, a lot of anxiety, a lot of stress” (SME). | “You know if you need medications, you need to draw up them before you go because it is really hard to do it in flight” (SME). | “We just work in a partnership really. So I think we complement each other. It depends on the circumstance” (SME). | “It is getting into the airport of xxxxxx, sliding them one stretcher to the next stretcher, putting them on a lifting device, getting them on a plane and then bringing them back down” (SME). | “If the woman is about to deliver we would communicate and ask air traffic control for a priority landing. If we want to descend to sea level, we would request this to the pilot through the crewmember” (SME). | “Calm under pressure, flexibility. Because you’re gonna be in a small space and have to use your skills in a small space that you used to use in a big room all with your stuff” (SME). |
| “You have to reassure the woman because they usually haven’t been in a smaller cabin space for a flight which is really noisy and cold” (SME). | “We always try to look at the worst-case scenario in all of our patients and what we are going to do if that scenario happens” (SME). | “I don’t think I have flown with a midwife twice. So you really have to communicate what are policies and procedures they are going to implement” (SME). | Her communications with team members were most of the time directed towards the anticipated issues and concerns. She often tried to convey her concern to other members even if it was only very slight (FO). | A” tight relationship with the flight nurses/ paramedics makes things so much safer. So you communicate with each other with trust” (SME). | Of course, you have to reassure the woman because they usually haven’t previously been exposed to such an uncomfortable, unfamiliar environment”(SME) |

SME- Subject matter expert, FO-Field observation

#### Task Management

Task management is a category of non-technical skills that health professionals use to manage and organise the various resources or tasks, and includes behaviours that demonstrate skills in (14):

- planning and preparing
- setting priorities
- maintaining standards
- identifying and utilising resources..

Planning, identifying and utilising resources (people, expertise, equipment, and time) before a maternity aeromedical transport mission comprises many elements, such as sourcing and organising an appropriate mode of transport or assessing the clinical risk of inflight delivery. A strong overriding message from all participants was that ‘planning for the worst and hoping for the best’ should be a focus for the transfer. In a maternity aeromedical transfer setting clinicians need to set their priorities depending on how much time is available, the condition of the pregnant woman, the weather, and what needs doing. Health professionals often have to decide which tasks are a priority, and respond appropriately. Protocols and guidelines are particularly important for multidisciplinary teams where the team members are not accustomed to working together, so adherence to best practice guidelines helps team members to understand what is expected of each other without limiting clinical judgment (74). This also involves strategies such as recording near misses and errors objectively (Table 4) (33, 75, 76)

#### Teamwork

It is crucial that multidisciplinary team members interact to facilitate a shared understanding of what is happening currently, and work towards a common goal in a team context (77–79). Teamwork includes the skill elements of supporting others, solving conflict, exchanging information, and coordinating activities (14). The subject matter experts interviewed for this study illustrated a number of ways of supporting others both practically, cognitively and emotionally in a constrained care environment like an air ambulance. This may include positive behaviours like providing backup to each other when needed, providing and accepting help, or sharing the duties among the flight nurse/paramedic and the midwife/doctor. The skill of solving problems/conflict is not only about managing the negative aspects which can lead to dysfunction, but can also involve positively framing situations and facilitating deeper analysis of a problem (57). Conflicts are often ‘potential’ rather than ‘actual’, and could be effectively solved in advance by negotiation. For example, an important behaviour emphasised during interviews was establishing roles clearly, and confirming responsibilities at the beginning of a transfer mission, to reduce the potential for misunderstanding and inaccurate or inappropriate expectations during flight. Another key teamwork behaviour considered helpful for avoiding conflict was creating an encouraging environment where team members can be assertive, but do so in a positive and respectful manner without drawing attention to potential mistakes through direct criticism (80). These behaviours can create a positive work environment based on trust and respect for each other to improve team coordination (Table 4).

#### Situational Awareness

Situational awareness is essentially about maintaining a dynamic awareness of an overall operational and clinical situation. It requires clinicians to maintain attention and concentration as well as using their working memory to process or make sense of information (14). For this reason, it is a nontechnical skill that is vulnerable to interruptions, distractions, fatigue, or task overload; as such it is a non-technical skill upon which many other skills rely (81). Situational awareness also relates to clinicians using their understanding of the current situation to anticipate or run through the possibilities of what may happen next, and to think about how problems might be overcome if they arise.

Interview participants considered the maternity transfer environment to be particularly ‘dynamic’ with respect to both clinical and aviation-related aspects (Table 4). An important factor here is that if the clinicians’ mental model is poor or incorrect, or they are relying too much on previous experience, their understanding of the current situation may be inadequate (14).

#### Communication with Team Members

Communication is a skill considered integral to all other non-technical skills (57). Exemplar behaviours for team communication in maternity aeromedical transfer setting in this study were determined by considering both patient care needs and operational/environmental factors. An additional challenge highlighted by study participants was that the team composition varies from mission to mission, and team members may meet each other for the *first time* when they arrive at the aircraft. Moreover, once on board team members cannot always easily communicate during the transfer due to aviation-related challenges such as aircraft noise or requirements to stay seated and restrained during large portions of the flight. These factors were carefully considered when formulating the exemplar behaviours for this non-technical skill category (Table 4).

#### Environment in the Cabin

As previously mentioned, the clinical and operational environment for a maternity aeromedical transfer has inherent physical challenges like vibration, noise, poor lighting, turbulence, and so forth. Nonetheless, despite clinical isolation and transport-related challenges, health professionals need to maintain an atmosphere of calm and professional care provision. The interview data and field observations illustrated how calm, confident, caring, and reassuring behaviours can make women feel they are in safe hands (Table 4). Other studies have similarly shown that the welfare of pregnant women is better when clinicians maintain a calm, caring approach while acknowledging any patient distress (36, 61, 73). This might be demonstrated by clinicians showing a willingness to answer questions and hear the woman’s concerns, or by explicit recognition and verbal acknowledgment that transfer to a tertiary hospital was unexpected and unplanned. Positive encouragement, assessment of wellbeing, and handing control back to the woman are therefore all exemplar behaviours illustrative of maintaining a calm and positive environment in the cabin.

### Evaluation of the NOAT framework

#### Face validity

The face validity of a non-technical skills framework is deeply intertwined with the strength of its development (82). For the NOAT framework this was mainly addressed during the development process by adopting the following strategies: systematically identifying those non-technical skills categories most closely reflecting those required in the maternity aeromedical transport setting (49), using data triangulation techniques to develop the behavioural exemplars, and reviewing and revising the framework during development to ensure the clarity and content of skills and behavioural descriptors (64).

The review of direct field observation notes and reflective notes re-confirmed the face validity of the NOAT prototype. Field observations of four maternity aeromedical transfers confirmed that all skill categories and behaviours represented by the framework (or the need for such skills) were able to be observed. The observations also indicated that some non-technical skills seemed to be of greater relevance for different clinical roles at different stages of the maternity air transfer mission. Overall, the findings from field testing indicated the NOAT prototype provided utility, salience, and applicability to assess non-technical performance of maternity aeromedical transfer teams. Finally, analysis of simulation-based training videos confirmed that the NOAT framework is sufficiently representative to capture all potential non-technical skills needed in a maternity aeromedical transfer setting (Figure 1).

**Figure 1:**
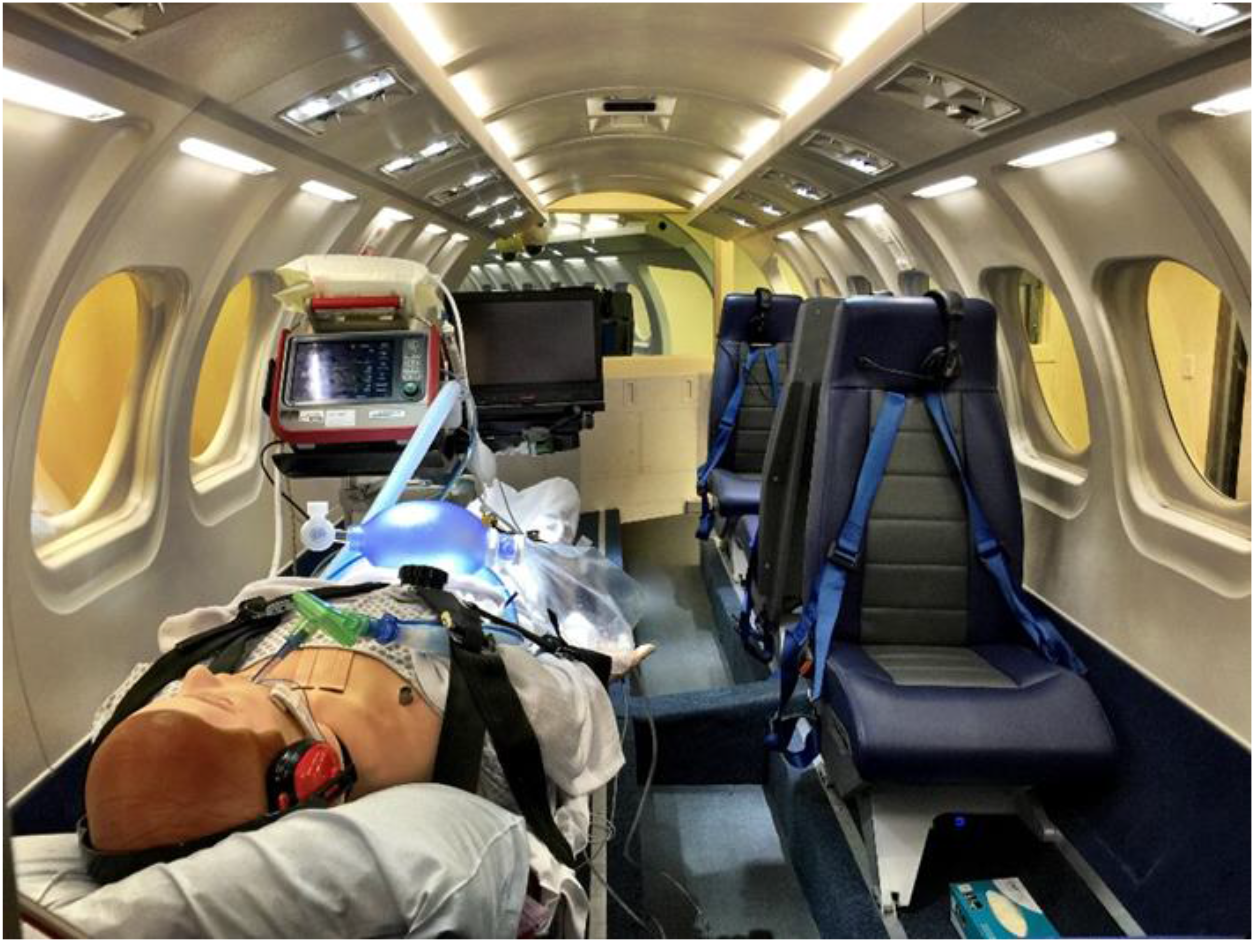
Flight simulator.

#### Content Validity

Representing all DHBs with a flight service in New Zealand, a total of seventeen fully complete anonymous surveys were returned from ten flight nurses, five flight midwives, and two paramedics; they had a median (IQR) aeromedical transport experience of eight (10–14) years. Of the 69 positive exemplar behaviours included in the online survey nine attained the maximum CVI of 1.0 (or 100%). For the rest of the 60 behavioural markers, 54 achieved the accepted level of CVI (75%) while six failed to achieve a CVI greater than or equal to 0.75; they were then reviewed and revised to finalise the NOAT framework (Table 5).

**Table 5:**
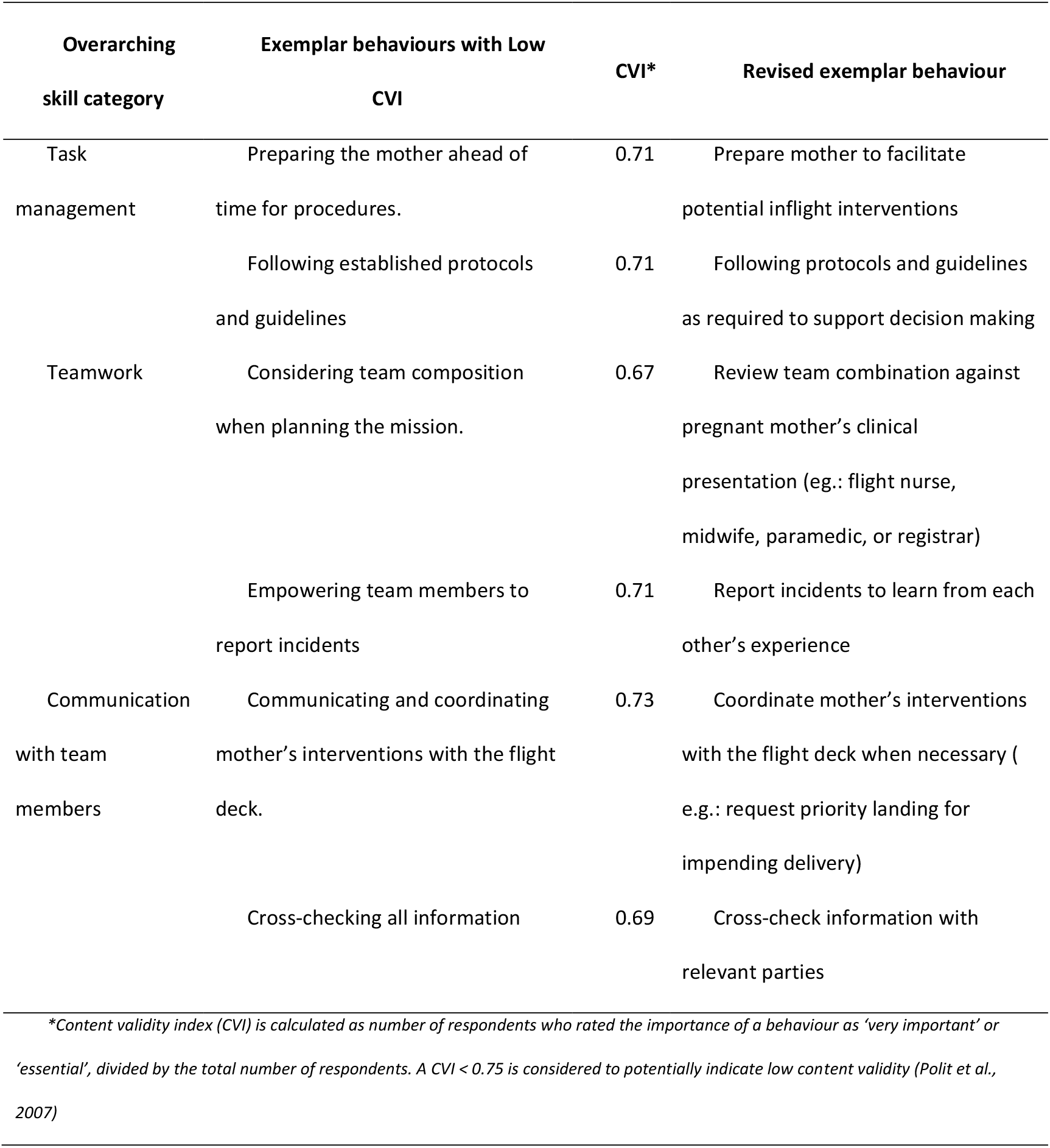
Behavioural descriptor modifications following Content Validity Index survey.

## Discussion

Comprehensive and consistent application of non-technical skills will enable clinicians to safely complete critical individual and team tasks in complex, high-risk, time-constrained, and dynamic clinical settings (14, 83). This can best be achieved by developing and applying non-technical skills frameworks appropriate for each clinical setting (57, 84, 85). In this study, we developed the NOAT behavioural assessment framework; a non-technical skills framework specific to multidisciplinary teams undertaking maternity aeromedical transfers.

Similar to other non-technical skill frameworks, NOAT was developed to provide a systematic means of describing and assessing the non-technical skills of maternity aeromedical transfer teams. It was based on the existing GAOTP framework but adapted for the maternity aeromedical transfer setting by triangulating data collected from a series of interviews with health professionals with subject matter expertise, direct observations in the field, and literature review. This type of transformational development approach is recommended when defining and refining the content of a behavioural marker system (64), and has been shown to be effective for eliciting knowledge on the non-technical skills required in specific clinical settings (57, 86, 87). Also, it provides a rapid, affordable, valid, and reliable method that provides impetus for the urgent development of new frameworks that can be applied widely across a wide range of clinical settings (64).

A behavioural framework must provide a means of recognising the execution of non-technical skills through observable behaviours (57) that can clearly indicate execution of the relevant skill (14). To make the relationships of non-technical skills and their behavioural markers clearer, the NOAT framework is arranged into a hierarchical structure with six main categories: communication with the pregnant woman and the support person/family, task management, teamwork, situational awareness, communication with team members, and the environment of the cabin. Related exemplar behavioural markers are grouped within these six categories and are written as action statements either describing directly observable behaviours, or behaviours from which the skills can reasonably be inferred.

Being able to evaluate the NOAT framework in a realistic air ambulance simulator was particularly useful as it was possible to create an atmosphere with all the inherent challenges of a maternity aeromedical transfer environment, including vibration, noise, limited space, turbulence, and poor lighting. It also provided an ideal and standardised opportunity to observe most of the unique non-technical skills transfer teams need to exercise in an aviation-based clinical setting. We were able to establish the face validity and content validity of the skill definitions and behavioural descriptors of the NOAT framework, suggesting it is a valid behavioural marker system capable of providing exemplar behaviours specific to the maternity air transfer setting.

While the behavioural markers of the NOAT framework are customised to maternity aeromedical transfer settings, it essentially comprises the main skill categories in the GOATP framework, which have been tested extensively for reliability, demonstrating good inter-rater reliability, internal consistency, and moderate test-retest reliability (62, 63). Confirmation of the reliability of skill categories from this comparative study can arguably be extrapolated to the reliability of skill categories of NOAT, as they do share largely similar constructs (88). A similar approach was taken to infer some of the findings of the reliability of GAOTP to that of AOTP, the framework which has the same skill categories across eighteen elements. However, it is advisable to conduct further testing on the reliability of the NOAT framework prior to using it as a standard evaluation instrument to measure the non-technical skills performance of maternity aeromedical transfer teams.

The multifaceted nature of maternity aeromedical transfer, where good quality non-technical skills during routine clinical care is critical to safety, places higher demands on health professionals in this clinical setting. An increased focus on the use of competency development in the exercise of non-technical skills therefore provides a crucial opportunity for clinicians and managers to enhance patient safety in the unique, dynamic, and potentially hazardous setting of high acuity maternity aeromedical transfers. The findings from this study suggest that the NOAT framework has high potential to be used as a training tool to improve non-technical skills in maternity aeromedical transfer teams, providing a common language and structure for feedback and debriefing purposes. Non-technical skills training could improve the maternity transfer team’s ability to recognise, operationalise, and apply non-technical skills into their clinical practice. The outcome of the implementation of such training could be investigated objectively and quantitatively using NOAT as a framework to measure the results. Incorporating the NOAT framework into professional training of maternity transfer teams will also enable health professionals to clearly understand each other’s role, to improve safe communication practices, to build joint understanding of clinical circumstances, and to make mutually agreed decisions. It could also be useful for service managers and policy developers to use to make informed decisions based on evidence, when making organisational reforms.

### Limitations

Developing and evaluating the NOAT framework involved clinicians and maternity air transfer services based in New Zealand only. However, every attempt was made to incorporate a more generalisable and global perspective by integrating all available evidence-based literature related to non-technical skills in obstetric and aeromedical clinical settings during development (36, 61, 85). The number of field observations was below what was initially planned due to logistic restraints imposed by the pandemic. Nevertheless, the transfers observed were diverse in terms of clinical details, flight team composition, time of day, overall mission time for the transfer, and the geographic origin of referral across New Zealand. We were therefore satisfied the observations and extensive field notes were representative of core maternity aeromedical retrieval practice.

## Conclusion

The NOAT framework developed in this study has the potential to significantly enhance the safety of pregnant women in a highly complex and dynamic maternity aeromedical transfer setting in a number of ways. Firstly, training and assessment of non-technical skills using NOAT could improve attitudes and standards of practice among clinicians in the maternity aeromedical clinical setting. This framework can also be applied to identify specific non-technical skills and their observable behaviours indicative of good and bad performance in this unique clinical setting, while the rating scale assigns a numerical value to the non-technical skill performance. Finally, encouraging results of the feasibility, content validity, and face validity of the NOAT framework suggest it can be used as a reference point to discuss and debrief non-technical skills, to scaffold structured teaching and training, and for service managers and policy developers to make evidenced-based decisions for organisational enhancements for maternity aeromedical transfer teams.

The development of Non-technical skills in Obstetric Aeromedical Transfers (NOAT) framework has also demonstrated that it is possible to transform non-technical skills frameworks originally designed for use in hospital settings to specialised, non-standard, out-of-hospital clinical settings such as high acuity maternity aeromedical transfers.

### Ethics approval

This study involved human participants and was approved by the University of Otago Human Ethics Committee (Health): H19/077. Locality approval to undertake this research study was also provided by District Health Boards (DHB) throughout New Zealand, and included specific Māori consultation in the respective regions. The study took place after obtaining informed consent from the participants. No data can identify individual participants.

## Data Availability

All data produced in the present study are available upon reasonable request to the authors

